# Constructing expert models of adolescent vaping behaviour: A mental models study

**DOI:** 10.64898/2026.10.26.26366558

**Authors:** Siv Hilde Berg, Kristine Stifjell, Kristin Benjaminsen Borch, Therese Haugdahl Nøst, Jo Røislien, Torkjel M Sandanger

## Abstract

Research on vaping prevention messages has predominantly examined adolescents’ perceptions of and responses and the research exploring how experts’ perspectives is limited. Expert knowledge is crucial for conceptualising the risk phenomenon and its key causal mechanisms, uncertainties, and issues that are relevant to address in evidence-based communication strategies. This study therefore explored how professionals and experts conceptualise and understand adolescent vaping behaviour, including its perceived causes and drivers.

Semi-structured interviews were conducted with a purposive sample of 12 experts from medical research, addiction science, public health prevention, and adolescent-oriented practice in Norway. Data collection and analysis were informed by the mental models approach to risk communication.

The analysis identified an expert-derived mental model/influence diagram of adolescent vaping behaviour. Across expert domains, vaping was understood as a multifactorial phenomenon emerging from interacting biological, psychological, and social factors operating across multiple system levels. The first theme describes vaping initiation as a socially embedded and often intermittent practice shaped by identity development, peer belonging, and low perceived harm. The second theme captures how vaping is enabled, sustained, and legitimised through interacting drivers operating across social, structural, commercial, and regulatory contexts. The third theme focuses on reinforcement processes, whereby nicotine dependence and emotional coping mechanisms contribute to more sustained use. According to these expert accounts, adolescent vaping is sustained and legitimised through broader processes of normalisation operating across multiple societal levels. The expert perspectives identified in this study suggest that comparative risk framings of e-cigarettes as less harmful than cigarettes, and the social functions of vaping within adolescent peer cultures, may be important considerations in the development of risk communication targeting adolescents.

## 1 Introduction

The use of e-cigarettes has increasingly become a topic of concern in public health, particularly in relation to adolescent use [1–2]. Concerns have been raised regarding nicotine dependence, smoking initiation, and the marketing of e-cigarettes to children and adolescents [1–4]. The World Health Organization has called for “urgent and strong decisive action” to prevent non-smokers from initiating e-cigarette use and developing nicotine dependence [1, 2]. Whereas proponents of a precautionary approach emphasise uncertainties and potential harms, others view e-cigarettes as a potential harm reduction tool for people who smoke combustible cigarettes [5]. Consequently, perspectives on the risks and appropriate risk communication of vaping vary across public health and scientific communities.

The existing health and risk communication literature regarding vaping prevention remains heavily oriented toward audience-centred approaches, particularly on how adolescents interpret, perceive, and respond to vaping prevention messages [6–7]. According to the mental models approach to risk communication, not only the perspectives of the target group are crucial for developing evidence-based communication strategies, but also those of the expert knowledge holders. Experts can pinpoint their construction of complexities, uncertainties, and causalities that are vital for a comprehensive understanding of vaping behaviour [8]. The current research on adolescent perceptions and message effects needs to be supplemented with research on expert knowledge and perspectives.

The mental models approach to risk communication outlined by Morgan et al. [8] provides a framework for mapping expert knowledge as causal systems, identifying key drivers, uncertainties, and relationships relevant for informed decision-making. This approach to systematically eliciting and representing expert knowledge is particularly valuable in a field where the evidence base is heterogeneous and rapidly evolving [9] as is currently the case for vaping research. According to Bruine de Bruin and Bostrom [10], expert mental models constitute an essential foundation for evidence-based communication because they provide a benchmark against which target audience understandings, knowledge gaps, and misconceptions can be assessed. The study seeks to answer the central research question: How do professionals and experts conceptualise and understand adolescent vaping behaviour, including its perceived causes and drivers?

## 2 Method

The present study applies a qualitative method designed as an individual interview study with Norwegian domain experts from medical research, drug science, public health prevention, and professionals working directly with adolescents.

The present study focuses on the first step of the mental models framework by eliciting expert knowledge to develop an expert mental model of adolescent vaping behaviour [8]. Similar expert-focused mental model studies have been conducted within other public health topics [8, 11–12]. Following this tradition, the present study uses in-depth qualitative interviews to elicit experts’ mental representations of vaping behaviour, including perceived causes, drivers, contributing factors, and their interrelationships.

### 2.1 Participants and sampling strategy

Expertise regarding vaping behaviour encompasses a range of interprofessional disciplines and practices across various levels of society. We therefore purposively sampled 12 experts and professionals, six males and six females, representing four domains: medical research, drug science research, public health prevention, and adolescent-oriented practice (hereafter referred to as “the experts”). The experts held diverse professional roles, including researchers, clinicians, public health practitioners, and frontline professionals working directly with adolescents.

To capture perspectives across system levels, the sample included experts operating at community, regional, and national levels. This multi-level composition enabled the study to explore how adolescent vaping is conceptualised across different institutional contexts, from everyday practice with youth to national public health and regulatory frameworks.

Participants who met the inclusion criteria were identified by the research team and recruited by invitation from the research group (SHB, KS or TMS). The sample size was guided by the concept of information power [13]. The interview provided rich data centred around a narrow research question, which supported a relatively small but information-rich sample. The experts, their professional roles and system levels are presented in Table 1.

**Table 1.** Sample characteristics.

| **Expertise domains** | **Professional roles** | **System levels** | **ID** |
|---|---|---|---|
| Medical research | Professor, oncology and tobacco-related health research | Regional academic and healthcare sector | 1 |
| Medical research | Senior researcher, toxicology | National public health research institution | 2 |
| Medical research | Professor, preventive medicine and epidemiology | Regional academic and healthcare sector | 3 |
| Drug science research | Senior researcher, sociology and nicotine research | National public health research institution | 4 |
| Drug science research | Researcher, sociology and nicotine research | National public health research institution | 5 |
| Public health prevention | Child welfare leader, youth outreach services | Community level | 6 |
| Public health prevention | Public health professional, health communication and prevention | National public health authority | 7 |
| Public health prevention | Professor, community medicine and health systems research | Regional healthcare sector | 8 |
| Public health prevention | Psychologist, child and adolescent mental health and prevention | Regional prevention and competence service | 9 |
| Adolescent-oriented practice | Youth support professional | Community level, lower secondary education | 10 |
| Adolescent-oriented practice | Youth support professional | Community level, lower secondary education | 11 |
| Adolescent-oriented practice | Service-user representative and youth support professional | Regional public welfare services | 12 |

### 2.2 Interviews

A semi-structured interview guide was developed for this project to explore expert understanding of vaping behaviour and risks, see the interview guide in appendix 1. The interview guide was used flexibly to adapt to each expert’s specialty. Nine interviews were conducted via Teams, digital video conferencing software, while three interviews were conducted in person with the informants. The interviews lasted on average 55 minutes (range 43-90 min). Interviews were audio recorded using *Nettskjema*, a GDPR compliant tool for managing data collection. SHB transcribed the interviews verbatim. The study was reported to and approved by Sikt - Norwegian Agency for Shared Services in Education and Research (Ref. 222521) in accordance with UiT’s procedures for the processing and storage of research data. All participants provided informed consent. The interviews were conducted between March 2025 and Nov 2026.

### 2.3 Context

This study was conducted in Norway, where e-cigarettes are subject to regulation [14–15]. In short, the sale of nicotine-containing e-liquids through domestic retailers is prohibited and flavours other than tobacco, such as spices, fruit or candy, are not permitted in e-cigarettes and e-liquids sold in Norway. Further, the minimum age for purchasing e-cigarettes and related products is 18 years, and private individuals are not permitted to import e-cigarettes or e-liquids from other countries. Vaping is prohibited in the same places where smoking is already banned, applying to both nicotine and non-nicotine e-cigarettes [16]. Although adolescent vapers should not have access to e-cigarettes, particularly nicotine-containing products, such products—including e-liquids and related equipment—are known to be purchased illegally [17].

The implementation of strict tobacco control policies in Norway, have contributed to a marked decline in cigarette smoking and reduced everyday exposure to smoking/tobacco among younger generations. However, recent Norwegian data suggest that vaping has increased among adolescents. In 2025, 13% of women and 6% of men aged 16–24 years reported occasional e-cigarette use, whereas daily use remained at approximately 1%. Among those who reported using e-cigarettes daily or occasionally in 2025, approximately 77% used nicotine-containing e-cigarettes, around 11% used nicotine-free e-cigarettes, and 12% did not know whether the e-cigarette they used contained nicotine [18].

### 2.4 Analysis

The expert interviews were analysed using inductive thematic analysis following Braun and Clarke’s [19] six-phase framework. First, SHB transcribed and read all interviews to become familiar with the data; KBB also reviewed the transcripts, and initial impressions were discussed within the research team. SHB then generated initial inductive codes. These were organised into a four-level hierarchical structure (individual, peer, social media, and regulation) to identify patterns and relationships, and reviewed by all co-authors in analytic workshops and online meetings.

Themes were subsequently reviewed and refined to ensure coherence and distinction, and a structured coding document was developed to validate them against the dataset. The final themes captured experts mental models of key drivers of vaping behaviour, outlined as interconnected biological, psychological, and social factors across system levels. In the final phase, results were written, illustrative extracts selected, and interpretations discussed among co-authors. The themes represent the informants’ understanding of vaping behaviour, and the influence diagram is an analytical construction and visualisation of the expert domain informants’ causal understandings of adolescent vaping behaviour.

The research team’s interdisciplinary background in public health, psychology, and health communication informed both the design and interpretation of the study. Reflexive discussions were conducted throughout the analytic process to critically assess how preconceptions may have shaped coding and interpretation.

## 3 Results

The analysis identified three interrelated themes capturing expert understandings of adolescent vaping. Across expert domains, adolescent vaping was understood as a multifactorial process emerging from interacting biological, psychological, and social factors operating across system levels. The first theme describes how the experts understood vaping initiation, and vaping as an intermittent phase-like and socially embedded practice within adolescent peer cultures, shaped by processes of identity formation, belonging, and low perceived harmfulness. The second theme captures how the experts understood adolescent vaping behaviour as being enabled, sustained, and legitimised through the interaction of multiple system-level drivers. The third theme focuses on processes of reinforcement, in which nicotine exposure and emotional coping mechanisms were understood by experts to contribute to more sustained use among a vulnerable subgroup of adolescents. Table 2 presents the thematic organisation of the findings.

**Table 2.** Themes and sub-themes describing experts understanding of adolescent vaping behaviour.

| Theme | Sub-themes |
|---|---|
| **Vaping as a phase of adolescence in a peer-driven social context** | • The occasional vaper<br>• A marker of group identity and belonging<br>• Risk trivialisation and knowledge gaps legitimising use |
| **Vaping as a low-threshold and systemically legitimised adolescent behaviour** | • Product appeal through social media<br>• Limited regulation and weak enforcement<br>• Relativised risk and systemic ambivalence |
| **Addiction reinforcing further vaping behaviour** | • Nicotine addiction and multiple nicotine product use<br>• The externalisation of emotional coping |

The influence diagram (Figure 1) is empirically grounded in the interview data and illustrates experts’ understanding of adolescent vaping as a multi-level process. The influence diagram visualises how experts understood the mechanisms forming vaping behaviour, in which the system conditions were understood to contribute to a broader process of normalisation, through which vaping was framed as acceptable and low-risk, shaping peer contexts and individual perceptions that drive initiation. Vaping initiation was furthermore understood as a socially embedded and intermittent practice, which may develop into reinforced use through nicotine exposure and emotional coping mechanisms. Individual vulnerability was understood to moderate the risk of addiction. Addiction was not understood as an essential driver of initiation, but rather as a consequence and a mechanism reinforcing continued vaping behaviour.

**Figure 1.**
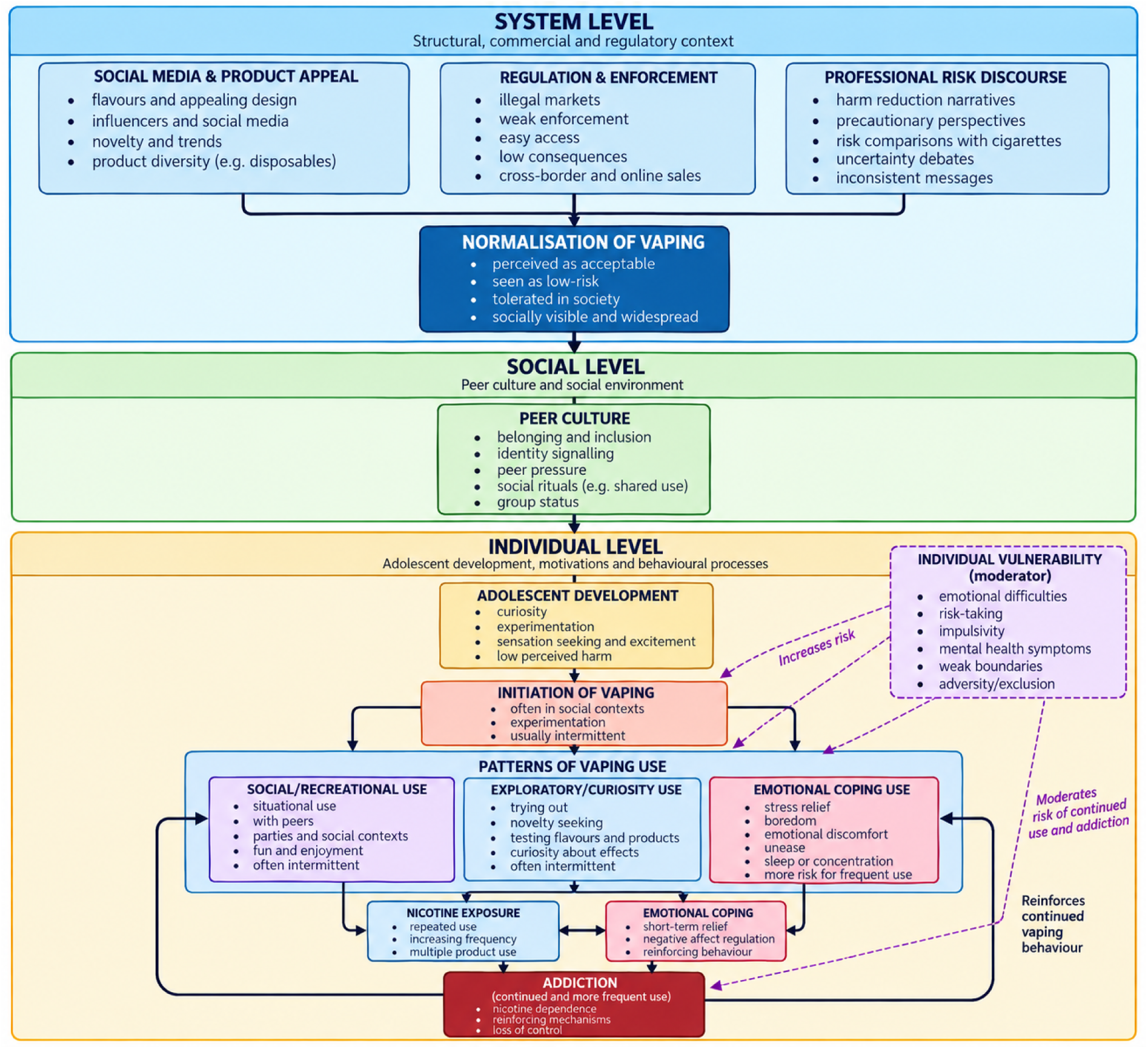
Influence diagram of expert’s understanding of adolescent vaping behaviour

### 3.1 Vaping as a phase of adolescent in a peer driven social context

As illustrated in the influence diagram (Figure 1), peer context and individual factors represent central pathways through which broader system-level conditions are translated into vaping initiation. This theme shows that experts conceptualised adolescent vaping as an intermittent and socially embedded practice driven by peers, and shaped by processes of group belonging, identity formation, and low perceived harmfulness. Experts described the largest group of adolescent vapers as *the occasional vaper*, as adolescents are inherently risk-seekers driven by behaviour in peer-oriented social contexts. The function of vaping was understood as a *marker of group identity and belonging*, in which *risk trivialisation legitimises use,* while *knowledge gaps* regarding potential risks further reinforce this perception.

#### 3.1.1 The occasional vaper

The experts conceptualised adolescence as a developmentally sensitive period characterised by heightened responsiveness to novelty, peer influence, and low-threshold risk-taking. Vaping activates these age-specific dynamics by offering an accessible way to experiment with boundary-testing behaviour that is perceived as low in severity in terms of perceived health effects (ID: 3, 6, 9, 10, 11).

Adding to the general developmental vulnerability of adolescence, experts described current cohorts as particularly exposed to social conditions that increase susceptibility to risk-oriented behaviour (ID: 6, 10,11). One social worker emphasises that lower secondary students are especially prone to substance use, framing this group as a generation characterised by increased norm-breaking (ID: 6). She linked this to post-pandemic changes, including greater visibility in public spaces and more frequent engagement in behaviours such as smoking. These developments were understood as rooted in disrupted socialisation, growing inequalities, and experiences of exclusion, which in turn drive young people to seek belonging in peer environments where norm-breaking functions as a marker of affiliation.

Across expert domains, vaping was conceptualised as an intermittent and largely transient phenomenon, particularly among eighth-grade students in the first year of lower secondary school (ID: 4, 5, 6, 9, 10, 11). The “everyday vaper,” or clear behavioural indicators of nicotine dependence, was described as relatively uncommon in lower secondary school contexts (ID: 4, 5, 6, 9, 10, 11). As one drug science researcher noted:

> “Most adolescent vaping seems to be transient and experimental. There is very little regular, daily use among adolescents. Instead, it appears to be something that happens in phases—it becomes a hype, and then it fades and they stop.” (ID: 4)

Several experts emphasised that vaping is often seen as losing its appeal as adolescents grow older and transition into tenth grade and upper secondary school (ID: 5, 10, 11). As one drug science researcher noted, *“This may indicate that attention is directed toward the playful or exploratory character of the behaviour”* (ID: 5). At the time of the interviews, this developmental pattern reinforced experts’ interpretation of vaping as primarily a phase-like phenomenon rather than a behaviour with high perceived addiction potential.

Although the typical adolescent vaper was conceptualised as an occasional social vaper across expert accounts, the phenomenon was also acknowledged as multifaceted, as described by a drug science researcher:

> “There is no single set of motivations, and the user group is not uniform. People use it for different reasons, engage in different practices, and face very different levels of risk. This makes prevention work difficult and raises questions about which strategies or ‘buttons’ to push.” (ID: 5).

Adolescents who develop stronger or more persistent dependence through vaping were understood to differ from the majority of experimental users. Across experts, these adolescents were characterised by additional risk factors, including limited boundaries at home, broader risk-taking behaviour, emotional regulation difficulties, and experiences of social exclusion (ID: 6, 9, 10). Experts emphasised that for this subgroup, vaping dependence often forms part of a wider pattern of vulnerability, rather than emerging from vaping alone.

#### 3.1.2 A marker of group identity and belonging

Adolescent vaping was consistently conceptualised by experts as a socially embedded practice that serves important identity-forming and relational functions within peer groups. Rather than being driven primarily by individual preferences or substance effects, vaping was framed as a collective activity through which adolescents negotiated belonging, status, and social identities associated with being “cool”. Several experts emphasised that vaping often unfolds within specific social rituals and contexts—such as school breaks, parties, or shared pauses, often in school toilets—where engaging in vaping legitimises participation in group activities and temporarily suspends other demands, giving the practice a ceremonial and contextual character (ID: 3, 4, 10, 11).

Experts further conceptualised vaping as a visible marker of identity. What product is used, how it is used, and with whom it is shared were understood to communicate signals about identity, taste, status and group affiliation, in ways comparable to clothing, style, or other symbolic markers. In this sense, vaping functions as a distinction marker, allowing adolescents to position themselves socially and to be read by others in intended ways (4), as described by the drug science researcher: *“Nicotine use, in general, contains an identity-forming element. The type of product used, the manner of use, and the social context in which it is used can all signal aspects of one’s identity to others”* (ID: 4).

Vaping was described as something adolescents explore together. Experts emphasised that initial engagement often occurs through friendships, where trying vaping collectively becomes a shared project that strengthens social bonds. This shared exploration—selecting flavours, devices, or experimenting with effects—was understood as enjoyable in itself, independent of substance content, and as a way of building companionship and mutual experience (ID: 4, 9).

Group pressure emerged as a central mechanism shaping vaping behaviour, although experts stressed that such pressure is rarely explicit. School-based professionals described situations in which adolescents participate in vaping not because they want to, but because refusing risks exclusion from the group. In unstable social environments, where peer groups are described as fluid and insecure, joining peers in vaping is described as a strategy to avoid being left alone, marginalised, or pushed out of social spaces (ID: 3, 10, 11, 12). Participation also created implicit expectations of loyalty, where being part of shared rule-breaking reduces the likelihood of being reported or excluded, further binding individuals to the group and lowering barriers to continued involvement (ID: 10). Not wanting to be perceived as an outsider therefore functions as a central driver of vaping behaviour:

> “I actually think there is a lot of influence operating here—when one person does it, others follow. It’s almost as if, if you don’t take part, you are seen as an outsider. I think that has a lot to do with it. If you feel unseen or unheard, that can trigger you to try it. And this doesn’t necessarily require belonging to a close-knit group—it can operate at the level of the whole school. If others are doing it, you do it too, and you are pushed into it even if it wasn’t something you had decided. It becomes a trigger, quite simply.” (ID: 12).

Experts also linked vaping as a group marker to broader changes in adolescents’ social environments, particularly the intensified role of digital platforms. Group chats, social media, and location-sharing technologies were described as amplifying feelings of comparison and exclusion by making social gatherings continuously visible. Within these contexts, adolescents were seen to navigate heightened pressure to be included, where not participating—or not being present—becomes immediately noticeable. Vaping was thus understood as one of several practices through which adolescents attempt to secure belonging and visibility in peer networks experienced as competitive and emotionally demanding (ID: 10, 11, 12).

#### 3.1.3 Risk trivialisation and knowledge gaps legitimising the use

Experts described adolescent intermittent vaping as categorically different from adolescent smoking and therefore less readily associated with addiction or long-term health risks. This normalisation of the behaviour, together with adolescents’ risk comparisons with high-risk and hazardous cigarettes, was further understood as reinforcing risk trivialisation. Risk trivialisation thus emerged as a driver both in adolescents’ understandings of vaping and, more broadly, in how vaping is framed in public discourse, which in turn contributes to the normalisation of the behaviour (ID: 4, 5, 10, 11).

Moreover, excitement and curiosity were repeatedly identified by experts as central psychological drivers of adolescent vaping and were framed as part of normative developmental processes (ID: 4, 5, 9, 10, 11). As one community psychologist explained, *“They vape out of curiosity. And they have a need to be engaged in something like that”* (ID: 9). Experts described vaping as a form of experimentation aligned with normative adolescent boundary-testing. Within their mental models of adolescent vaping behaviour, experimental engagement with a product perceived as low harm, yet with a clear risk element, was understood as a key driver of occasional use, as illustrated by this statement from a school-based professional in lower secondary school:*“It is a form of experimentation for adolescents. It is exploratory. It is about excitement. It is somewhat illegal, but not entirely… It is not narcotics. It is not really crossing the line in a major way.”* (ID: 10)

In addition, experts highlighted substantial gaps in adolescents’ knowledge about vaping products and associated health risks as a contributing factor to low perceived harm. The toxicologist emphasised that vaping was not a uniform product; risk was described as depending on what is used, how it is used, and what substances are added to the device (ID: 2). Experts further emphasised that adolescents may be unaware of whether and how much the products they use contain nicotine, and that devices may include substances such as THC or CBD (11). Moreover, experts emphasised that adolescents lack knowledge about what they are vaping. Even regulatory authorities are described as lacking a complete overview of permissible substances in e-cigarettes, as the product category is understood to remain largely unregulated (ID: 2, 4, 5, 7).

### 3.2 Vaping as a low-threshold and systemically legitimised adolescent behaviour

As illustrated in the influence diagram (Figure 1), experts conceptualised system-level conditions as contributing to adolescent vaping indirectly through broader processes of normalisation. This theme captures how product characteristics, social media dynamics, regulatory gaps, and frameworks of relativised risk collectively shape a context in which vaping is perceived as acceptable, accessible, and low-risk. Within experts’ mental models, *product appeal through social media* lowers the threshold for initial use by framing vaping as an attractive, novel, and experimental part of peer practices. These dynamics are further supported by *limited regulation and weak enforcement* and *competing risk logics*, where vaping is downplayed by a large group of experts through comparison with smoking and deprioritised within prevention systems.

#### 3.2.1 Product appeal through social media

Experts described product characteristics of e-cigarettes as central drivers of adolescents’ experimentation with vaping. Product features and their appeal through social media frame vaping as playful and exploratory, contributing to low perceived harm and the normalisation of vaping among adolescents.

The frequent introduction of new devices was understood to generate immediate curiosity, particularly when products appear more technologically advanced or visually distinctive than earlier versions (ID: 4). As one drug science expert noted:

> “New products are constantly being introduced, and some of them may seem cooler than the old ones. That creates curiosity about products with lots of extra features, like touch screens, Bluetooth connectivity, and all sorts of things (ID: 4).

He further perceived disposable e-cigarettes as especially appealing, as they are understood to be accessible, novel, and well suited to short-term experimentation rather than sustained use (ID: 4).

Flavours were similarly emphasised as a major source of appeal, lowering the threshold for experimentation and reinforcing vaping as an activity driven by excitement and sensory curiosity rather than by a deliberate pursuit of nicotine effects (ID: 2, 4).

Product appeals through social media were further perceived as powerful influences on adolescents’ vaping behaviour, particularly when messages were repeated by influencers or adults. Combined with adolescents’ limited source criticism, these appeals reinforce misconceptions about harm and contribute to the normalisation of risk (ID: 10).

#### 3.2.2 Limited regulation and weak enforcement

Adolescent vaping was understood to occur in a context characterised by limited regulation and weak enforcement across multiple arenas. Experts described a lack of consequences from parents, schools, and authorities as contributing to a permissive environment in which vaping can continue largely unchallenged (ID: 8, 10, 11).

Adolescent vaping was frequently described as taking place within illegal and largely unregulated markets, where products are often obtained through social media platforms such as Snapchat, where adolescents connect with sellers (ID: 4, 5, 8, 10, 11). As one school-based professional noted:

> “It’s Snapchat. If you don’t know where to buy, if you haven’t done it before and are going to start buying, you just search for ‘Puff’. Then a lot of different sellers come up that you can buy from.” (ID: 10)

Experts also emphasised the lack of effective regulation of illegal sales through social media platforms. This was framed as particularly problematic given that very young adolescents can arrange transactions with strangers and engage in illicit purchasing practices within potentially risky drug markets (ID: 4, 5, 8, 10, 11, 12). This regulatory gap is further facilitated by a lack of system-oriented prevention efforts, with insufficient coordination between prevention, regulation, and enforcement (ID: 8).

Moreover, school-based experts reported a lack of enforcement of rules by both parents and schools. Parents are frequently described as unaware of vaping products and practices, with limited knowledge of what devices look like or what to look for. This lack of parental awareness reduces the likelihood of detection and consequences at home, thereby weakening preventive efforts (ID: 10, 11). Vaping is also described as largely absent from formal drug prevention strategies at the school level, which further contributes to its normalisation (ID: 10, 11). Unlike smoking, vaping was described as difficult to detect and regulate in school settings due to the absence of smoke and persistent smell, often masked by perfume. This practical invisibility makes it challenging for staff to identify individual users or impose consequences, leading to uncertainty and hesitancy in enforcement.

Within experts’ mental models, this absence of clear responses in adolescents’ environments was understood to signal a form of tacit acceptance, thereby weakening the perceived seriousness of vaping as a risk behaviour (ID: 10, 11). The combination of easy access, novelty, and illegality, together with weak enforcement, reinforces the experimental and risk-seeking aspects of vaping behaviour (ID: 4, 5, 8, 10, 11).

#### 3.2.3 Competing risk logics

Adolescent vaping behaviour was framed as a comparatively minor concern by some experts, and as a significant risk by others. This disagreement revealed a structural tension between harm-reduction and precautionary approaches to risk, which shapes the public discourse of vaping risk and thereby also its prevention efforts.

The drug science experts consistently assessed vaping in comparison to smoking, as well as in relation to other forms of substance use and broader public health challenges. They did not understand vaping as a major public health problem comparable to smoking, due to both lower assumed health risks and significantly lower prevalence (ID: 4, 5). Smoking was understood to occupy a fundamentally different risk category, often characterised as *“in a league of its own”* (ID: 5). In contrast, vaping was understood as a lower-impact phenomenon at the population level, particularly given predominantly occasional use among adolescents (5). As one drug science researcher explained:

> “If we look at whether vaping can become a public health problem in the same way as smoking, it never will. That has to do with both lower risk and much lower prevalence” (ID: 4).

The drug science researcher explained how within prevention work, professional experience with severe substance use and tobacco-related disease shaped a relatively high threshold for concern, which in turn influenced how vaping was evaluated and prioritised (ID: 5). Within this comparative risk perspective, school-based professional described that adolescent vaping was often interpreted in school contexts as less urgent than other problems, thereby reducing its salience in prevention efforts (ID: 10, 11). Some of the experts argued for a pragmatic approach to risk, arguing that adolescent is inherently risk seeking, and having vaping as a low-risk alternative may be a safer option, than adolescents choosing other more harmful substances (ID: 4, 5, 10, 11).

At the same time, other medical experts were clear that lower relative risk should not be conflated with safety. These experts emphasised a precautionary stance towards adolescent health. They argued that uncertainty regarding long-term health effects, early nicotine exposure during developmental periods, and the ethical implications of normalising experimentation among young people necessitate a more restrictive and cautious approach, irrespective of relative risk comparisons (ID: 2, 3, 8). Others argued that the risks associated with e-cigarettes were substantial, particularly in light of concerns regarding nicotine addiction and adolescent brain development. They perceived the availability of e-cigarettes as a smoking cessation tool as coming at the expense of children and young people, whose health and neurodevelopment were being placed at risk (ID:3).

A central point of disagreement concerned how uncertainty about long-term health effects ought to be weighed in relation to adolescents. Some medical experts emphasised that the evidence base for vaping remains incomplete, particularly with regard to long-term outcomes such as cancer, cardiovascular disease, and respiratory harm (1, 2). This uncertainty is itself framed as a significant risk, especially when products are used during critical developmental periods. The toxicologist critically questioned the widespread use of comparative risk arguments:*“It’s become completely wrong to argue that it’s ‘not so dangerous’ just because it’s less harmful than smoking. These are products that carry a health risk, and we don’t know the full consequences yet—especially when you start experimenting with 13-year-olds”* (ID: 2). A researcher on cancer and smoking risk, argued that it was naïve to wait for definitive evidence that e-cigarettes cause cancer, given the existing knowledge that many of the carcinogenic substances found in e-cigarette aerosols are already known to cause cancer (ID: 3). Within this precautionary approach to risk, the focus for some experts shifted from individual biomedical harm to broader normative concerns. The key question is not only how harmful vaping is. Several experts raised concerns about vaping as a potential gateway to broader substance-use trajectories, particularly among adolescents already oriented towards risk-taking (ID: 6, 9, 12). Others highlighted that even if vaping does not lead to substantial disease burden, its normalisation may alter norms around substance use and rule-breaking during adolescence, with possible downstream effects (ID: 8).

### 3.3 Addiction reinforcing further vaping behaviour

As illustrated in the influence diagram (Figure 1), this theme captures the processes through which adolescent vaping, following initiation, may develop into reinforced and more sustained patterns of use over time. Experts described how nicotine exposure and emotional coping functions contribute to continued use, positioning addiction not as a primary driver of behaviour, but as a consequence of prior engagement that reinforces further vaping.

In experts’ mental models, continued use was shaped through *nicotine addiction and multiple nicotine product use*, as well as through *the externalisation of emotional coping*, where vaping becomes a habitual strategy for managing everyday emotions.

#### 3.3.1 Nicotine addiction and multiple nicotine product use

Nicotine addiction was positioned as a mechanism that can reinforce and stabilize vaping behaviour over time, rather than as a primary driver of initiation. Early vaping among adolescents without prior experience with cigarettes or snus is not understood to be nicotine-driven but motivated by other psychological and social factors. However, the drug science experts emphasized that once nicotine exposure becomes more sustained, the risk of dependence increase (ID: 4, 5).

Nicotine was described as a stimulant that triggers neurobiological processes involving dopamine, serotonin, and adrenaline, producing pleasurable yet activating effects such as increased alertness and a perceived “kick” or boost and temporary cognitive effects, including enhanced attention and memory (ID: 3, 4, 9). While these effects are not understood as central motivations for initial experimentation, experts described them as becoming more salient with more regular use, thereby reinforcing continued vaping behaviour (ID: 4, 9).

Drug science experts further stressed that vaping should not be understood as an isolated behaviour, but as part of a broader pattern of nicotine use that frequently overlaps with snus and cigarette smoking. Within this understanding, co-use of nicotine products was seen as substantially increasing the risk of cigarette smoking and nicotine addiction. Repeated nicotine exposure is therefore understood as increasing the likelihood of continued vaping and, for a subset of adolescents, the development of dependence (ID: 5).

#### 3.3.2 The externalisation of emotional coping

Complementing biological perspectives, the some of the experts framed addiction as extending beyond nicotine’s neurobiological effects, emphasizing its social and psychological function in regulating everyday emotions (ID: 9, 3). Within this account, vaping was understood by the community psychologist to serve as a form of emotional coping—helping manage boredom, unease and restlessness, sadness, or discomfort—regardless of nicotine content (ID: 9).

> If you’re not feeling well, it may be that you eat too much, become addicted to exercise, or start vaping. Vaping then becomes a kind of comfort. And then we find comfort in other things too, and it turns into an addiction—it becomes a go-to. Instead of dealing with life, it becomes a go-to. Something that sort of keeps you afloat (ID: 9).

In this perspective, addiction was described as emerging when individuals become unable to cope with ordinary emotions, with vaping serving an emotion regulation function as a short-term relief at the expense of interfering with normal reward processes and emotional regulation.

## 4 Discussion

The findings show that experts conceptualise adolescent vaping behaviour as a multi-causal and dynamic process operating across interacting system levels. Vaping is positioned as a socially meaningful group practice driven by affiliation, identity signalling, ritualisation, shared exploration, peer pressure, and performative status, which is an acceptable, normalized and tolerable risk by both adolescents and their immediate social environment. Within this understanding, vaping is less about individual risk-taking and more as a relational strategy for managing belonging and social vulnerability within contemporary adolescent peer cultures. However, while these proximal drivers are primarily located within adolescent peer cultures, these behaviours are shaped and enabled by broader structural conditions. In particular, limited regulation of illegal products, the pervasive role of social media promotion, and a dominant risk discourse are highlighted as the reason why vaping can emerge, spread, and become normalized due to the framing of vaping as relatively low-risk compared to smoking.

These findings align with previous research showing that adolescents’ perceptions of vaping are strongly shaped by social norms and exposure to marketing, access and e-cigarette use among peers [6, 20–22]. However, whereas previous research has primarily focused on individual-level determinants and social influences [6–7], the present study suggests that the normalisation process is not confined to adolescent peer cultures alone, but also confined to the way e-cigarettes are marketed and regulated in the society, along with the public discourse of vaping risk.

The findings suggest that normalisation operates not only within adolescent peer cultures but also within professional and experts’ understandings of vaping risk. While several experts identified risk trivialisation as a mechanism contributing to vaping uptake, some experts simultaneously framed vaping as a relatively minor concern when compared to smoking. This reflects competing risk logics within expert discourse itself, where harm-reduction perspectives may unintentionally contribute to broader processes of normalisation. This highlights the importance of considering how macro-level contexts, social environments, and individual processes interact over time to produce risk behaviour. Future research should therefore move beyond individual-reductionist models and examine how structural, social, and professional understandings of risk interact over time to shape this behaviour [23].

This tension between precautionary and harm-minimization logics in how experts understand adolescent vaping behaviour has been identified as a conflict in the e-cigarette debate [5]. In this study, experts adopting a precautionary perspective emphasise absolute risk and the uncertainty surrounding long-term health effects, which holds that a lack of scientific certainty should not delay preventive action. In contrast, experts drawing on harm-minimization logic emphasise a relative risk perspective and the minimisation of overall harm, even in the presence of uncertainty [5]. Within this framework, risk behaviours are evaluated in relation to more harmful alternatives, and the aim is to reduce overall harm rather than eliminate risk entirely. Accordingly, these experts emphasise relative risk, often comparing vaping to more harmful behaviours such as cigarette smoking.

This internal tension within the expert panel establishes an analytically significant finding. Mental models do not necessarily display agreement across experts’ accounts, which are in line with other expert mental models’ studies [11]. How these accounts should inform a communication strategy is therefore also a normative judgement about how risk and uncertainty should be managed.

The evidence on risk communication is conflicting in how to approach expert disagreement, but studies indicate that if this disagreement is reproduced in the public health messaging, professional practice, and media coverage, it may increase perceived uncertainty and weaken the clarity of public health messaging and affect risk perception and trusted sources [24]. Moreover, when the public is exposed to conflicting messages and feels that authorities are “downplaying” information, or that they are not value-neutral in their presentation of risk, people often choose to trust information from other sources [25]. From a mental models perspective, the findings highlight comparative risk framings, scientific uncertainties, and the perceived social functions of vaping as potentially important topics for risk communication. These factors were identified by experts as influencing how vaping is understood, accepted, and normalised among adolescents.

Whereas recent consensus efforts have focused on synthesising evidence regarding the health effects of e-cigarettes and developing recommendations for practice [26], the present study contributes a complementary perspective by mapping how experts conceptualise the causal processes underlying adolescent vaping behaviour. Rather than identifying consensus alone, the study highlights both shared assumptions and competing risk logics within expert understandings and integrates these into a systems-oriented model of vaping initiation and continuation.

Furthermore, the present findings suggest that some drug science experts have a causal understanding of vaping behaviour as a temporally structured process, where initiation is socially embedded, while addiction emerges later as a reinforcing mechanism for continued use. This perspective contrasts with dominant biomedical framings, where nicotine dependence is typically treated as the central causal mechanism underlying use [4], in particular because dependence may emerge relatively early in the trajectory of use [27].

Although, this study only explores the perception of causal relationships, this conceptualization of vaping addiction may provide insight into why prevention messages focusing on addiction often show limited effectiveness among adolescents [7, 28]. If addiction is not a key driver, messages focusing on addiction and nicotine may fail, as they don’t address the motivational drivers of vaping initiation among adolescents. Messages related to “vaping causes nicotine dependence”, “this is dangerous for the brain” may not be relevant for the occasional and experimental vaper as they don’t experience to be driven by addiction. It’s the identity, the belonging social positioning and the hype, the messages need to address to reach this target group. This is supported by Gerson et al. [29] who argue that because adolescents may initiate or continue e-cigarette use as a way of coping with psychosocial stressors, prevention programmes should incorporate mental health education and adaptive coping skills to address these underlying risk factors for vaping initiation and addiction. Systematic reviews support that effective elements of school health promotion across behavioural domains, including substance abuse address social influences and norms, alongside with cognitive-behavioral skills to enhance self-regulation and coping skills [30].

The systematic review by Wu et al. [6] supports the importance of vaping status in targeting messages, as non-vapers have different risk beliefs and policy support of e-cigarettes than vapers. This study underscores that the primary drivers of vaping may differ among non-vapers, occasional vapers, and nicotine-dependent vapers, as well as across age groups within the broad age range during which children and adolescents initiate vaping. Consequently, prevention and communication strategies will require tailored messages for different target groups.

### 4.1 Limitations

This study employed a qualitative approach to analyzing how experts within different fields of expertise view the underlying causal mechanisms of this highly complex phenomenon. As the study consists of experts’ subjective perspectives on the topic, the presented findings aim to provide insights rather than generalisations. Furthermore, the assessments reflect the state of knowledge at the time of the interviews and should be regarded as a snapshot of expert perspectives in a field where the evidence base continues to develop.

The recruitment followed a purposive sampling strategy to ensure that participants would hold a wide range of expert backgrounds, including both research scientists and practitioners, on different system levels, thus providing a multi-systemic view. In similar research, a sample of 11-13 experts is often considered sufficient to allow for a rich understanding of the topic under study – here the potential mechanisms underlying adolescents vaping behaviour as viewed by experts in the field [11]. However – as our study has uncovered – vaping in adolescents as understood by experts is a highly complex phenomenon, and a larger sample or other fields of expertise might have revealed an even larger breadth of understanding of drivers and causes of vaping behaviour. Future studies with larger and more varied samples are needed to further explore the results from the present study.

## 5 Conclusion

Experts across domains conceptualised adolescent vaping as a dynamic and multifactorial phenomenon emerging from interacting biological, psychological, social, and structural influences operating across multiple system levels. The findings suggest that prevention and risk communication efforts may benefit from considering these multiple levels of influence rather than focusing solely on individual behaviour and risk knowledge. In particular, the expert accounts highlight the importance of social pathways of initiation and the social meanings and functions associated with vaping within adolescent peer cultures.

Furthermore, the prominence of comparative risk framings, particularly perceptions of vaping as less harmful than cigarette smoking, suggests that future risk communication may benefit from considering how absolute risks, scientific uncertainties, and the social acceptability of vaping are communicated. Together, these findings identify several factors that may be relevant to consider in the development of risk communication about adolescent vaping.

## Data Availability

The minimal dataset supporting the conclusions of this study, consisting of anonymised codes and illustrative quotations, is available from the University of Stavanger upon reasonable request to the corresponding author. Full interview transcripts are not publicly available due to ethical and confidentiality considerations, as participants were recruited as subject-matter experts and may be identifiable from contextual information contained in the interview.

## Acknowledgements

We would like to acknowledge the URØYK research team for their constructive feedback on the manuscript.

## Author contributions

SHB conceived the study, collected the data, conducted the interviews, led the analysis, and drafted the manuscript. KS contributed to participant recruitment, data collection, analysis, and manuscript development. KBB contributed to participant recruitment, analysis, and manuscript development. JR, THN and THS contributed to manuscript revision and critical review. All authors approved the final version of the manuscript. KBB, JR, THN, and TMS contributed to funding acquisition.

## Appendix 1

### Interview Guide for Expert Panel: Individual Interviews

This interview is part of research on youth vaping within the Sunne Valg project at the University of Tromsø. You are invited because you have different expertise that is important for informing how we design interventions to prevent youth from starting to vape. There are no right or wrong answers, and I do not have a specific answer I am looking for. Your experiences, assessments, and viewpoints are what matter here. I will use a Nettskjema app with a dictaphone, which also records audio. You consent to the storage of personal data via Nettskjema. The audio files will be transcribed into text, which will be stored in a password-protected area. Personal data will be anonymised when the study is published. No one other than researchers involved in the project will have access to the collected data. No health information about you will be collected in these interviews.

### Background

Can you introduce yourself, your professional background, and your area of expertise in relation to vaping and health communication?

### Causes

1. Why do youth in Norway vape?

- Are there factors that make some youth more susceptible to starting vaping?
- Social causes
- Psychological causes
- Societal causes
- The significance of legal aspects, regulation

### Risks

1. What are the consequences of vaping for youth?
2. Is youth vaping a significant risk in Norwegian society, or is it a minor risk? **Cause-Effect**
3. What is it about e-cigarettes that makes them harmful to health?
4. Can you explain how e-cigarettes cause harm? **Key Messages**
5. What is important to communicate to prevent youth from starting to vape?
6. What measures are important to prevent youth from starting to vape? **Communication Form**
7. What form should communication take when aimed at youth to prevent vaping?

- Which channels (social media, posters, news reports…)?
- What form (e.g., narrative style or facts, emotional or negative valence (focus on risk or focus on help and support))?
- How to reach out?

**Finally**, is there anything we haven’t discussed related to communication aimed at youth and vaping?

